# The Brain Imaging and Neurophysiology Database: BINDing multimodal neural data into a large-scale repository

**DOI:** 10.1101/2025.10.01.25337054

**Authors:** Charlotte Maschke, Peter Hadar, Yicheng Zhang, Jian Li, Gauri Ganjoo, Andrew Hoopes, Alessandro Guazzo, Aditya Gupta, Manohar Ghanta, Bruce Nearing, Christine Tsien Silvers, Bharath Gunapati, Robert Thomas, Jennifer A. Kim, Shibani S. Mukerji, Adrian Dalca, Sahar Zafar, Alice D. Lam, Emmanuel Mignot, M Brandon Westover

## Abstract

The Brain Imaging and Neurophysiology Database (BIND) represents one of the largest multi-institutional, multimodal, clinical neuroimaging repositories, comprising 1.8 million brain scans from 38,945 patients, linked to neurophysiological recordings. This comprehensive dataset addresses critical limitations in neuroimaging research by providing unprecedented scale and diversity across pathologies and health. BIND integrates de-identified data from Massachusetts General Hospital, Brigham and Women’s Hospital, and Stanford University, including 1,723,699 MRI scans (1.5 Tesla, 3 Tesla, and 7 Tesla), 54,137 CT scans, 5,093 PET scans, and 526 SPECT scans, converted to standardized NIfTI format following BIDS organization. The database spans the full age spectrum (newborn to 106 years) and encompasses diverse neurological conditions alongside healthy patients. We deployed Bio-Medical Large Language Models to extract structured clinical metadata from 84,960 brain-related reports, categorizing findings into standardized pathology classifications. All imaging data are linked to previously published EEG and polysomnography recordings from the Harvard Electroencephalography Database, enabling unprecedented multimodal analyses. BIND is freely accessible for academic research through the Brain Data Science Platform (https://bdsp.io/). This resource facilitates large-scale neuroimaging studies, machine learning applications, and multimodal brain research to accelerate discoveries in clinical neuroscience.

## 2 Background & Summary

Research on brain disease has benefited greatly from advances in and widespread adoption of neuroimaging for evaluation of neurologic disease.^1–3^ Although computed tomography (CT) of the head is the most common method for fast screening for gross abnormalities, magnetic resonance imaging (MRI) scans have unlocked the deep structures of the brain and serve as an essential adjuvant to the neurologic exam.^4^

Over the last decade, neuroinformatics has also witnessed significant growth.^2,5–7^ Enabled by advances in computer science, researchers have developed innovative computational methods to unlock the full potential of neuroimaging data.^8^ Today, machine learning techniques and computational neuroimaging analyses have demonstrated parity and superiority to human neuroradiologists.^9,10^ Subtle findings, which can only be detected with sophisticated computerized analysis of neuroimaging data in neurologic contexts including epilepsy, stroke, multiple sclerosis, traumatic brain injury, and dementia, yield novel insights which could not only revolutionize our understanding of neurological conditions but also contribute significantly to the advancement of human neuroscience as a whole.^11–15^

However, these advances have been tempered by small, siloed, narrow, and incomplete datasets that are collated at a limited number of institutions. Large datasets are needed to account for the vast range of diverse neurological conditions and significant inter-rater variability. While many groups have sought large databases, those databases are often dedicated to specific disorders and do not incorporate the natural variability seen in clinical neurologic care. Sharing data resources is crucial for driving innovation in neuroscience and development of clinical applications.^8^ In pursuit of our goal for open and shareable human neuroscience, our group has focused on collecting, collating, and analyzing large neurologic datasets that are made freely available to the research community.^16^

Here, we present the Brain Imaging and Neurophysiology Database (BIND), a large-scale database of multimodal imaging data comprising over 1.5 million scans from almost 39 thousand (n = 38,942) patients. Imaging data were obtained from patients who underwent clinical electroencephalogram (EEG) or polysomnogram (PSG) tests over the past 30 years. These include a wide variety of pathologies as well as normal findings over all age ranges, from newborns to the elderly. Available modalities include MRI, Positron Emission Tomography (PET), Single Photon Emission Computed Tomography (SPECT), and CT. MRI data were collected at different field strengths (1.5, 3, and 7 Tesla). Depending on the recording protocol available images are composed of structural, diffusion, perfusion, and functional sequences. The corresponding EEG and PSG data were previously published by our group and can be found in the Harvard Electroencephalography Database (https://bdsp.io/content/harvard-eeg-db/4.1/).16 An added benefit of this dataset, and of the BDSP platform, is that these data can be linked to additional neurologic testing (EEG, PSG) and electronic health records data. Data from all modalities are combined on the Brain Data Science Platform (BDSP; https://bdsp.io/), a freely available and easily accessible website that allows researchers to upload and share data with others and to download other data for research or clinical purposes. By providing these data to the research community, we aim to accelerate discoveries in the field of neuroscience and to unlock new avenues for research and the development of innovative clinical applications that can improve patient care and health outcomes.

## 3 Methods

### 3.1 Data Acquisition

The imaging dataset consists of clinical brain imaging scans acquired from three hospitals: Massachusetts General Hospital and Brigham and Women’s Hospital (collectively referred to as Mass General Brigham, MGB, labeled as I-1001) and Stanford University (labeled as I-1004). Scans were identified retrospectively from IRB-approved chart review under protocols approved by the BIDMC IRB (protocols #2022P000481, #2022P000417) and MGB IRB (protocol #2013P001024), which provided a waiver of consent for retrospective data analysis; no prospective data acquisition or participant recruitment was performed. Data from the MGB cohort include all patients who had undergone brain imaging in addition to EEG (routine or long-term monitoring). Data from the Stanford subset include all patients who had undergone brain imaging in addition to PSG testing.

### 3.2 Data De-identification

Data were de-identified by the corresponding de-identification services at Mass General Brigham (‘Medical Imaging Data As A Service’ (MIDAS)) and Stanford (Stanford Medicine Research Data Repository (STARR)). Demographics and imaging metadata were de-identified per each institution’s imaging service, and all data were date-shifted to further comply with HIPAA Safe Harbor de-identification standards. Free text of clinical reports were anonymized using automated de-identification software.^17^ To avoid identifiable information in the images themselves, all images were passed through the Tesseract optical character recognition (OCR) engine.^18^ Detected sequences with identifiable information were removed from the dataset. To further enhance security, the data are available via “controlled access”: all individuals seeking access to the data are required to sign a mandatory Data Use Agreement, which includes strict terms and conditions, and to provide proof of CITI training (https://about.citiprogram.org/). These agreements prohibit attempting to reidentify individual records or further sharing of the data. For more information, visit bdsp.io (https://bdsp.io/).

### 3.3 Computational Processing

All imaging data were converted from the Digital Imaging and Communications in Medicine (DICOM) format to the Neuroimaging Informatics Technology Initiative (NIfTI) format, using the software dcm2niix. The extraction of sequence names and clinical metadata was performed using custom software, described below.

#### 3.3.1 Sequence Names

To standardize MRI sequence naming across heterogeneous datasets, a metadata-driven approach was implemented based on acquisition parameters extracted from de-identified DICOM headers. Specifically, the following DICOM tags were used: Image Type (0008, 0008), Scanning Sequence (0018, 0020), Sequence Variant (0018, 0021), MR Acquisition Type (0018, 0023), Echo Time (TE) (0018, 0081), Repetition Time (TR) (0018, 0080), Inversion Time (TI) (0018, 0082), Flip Angle (0018, 1314), Diffusion B Value (bval) (0018, 9087), and Echo Train Length (0018, 0091). Using a combination of parameter-guided thresholds (based on MRI physics knowledge) and unsupervised clustering of these parameters using Gaussian Mixture Models on the I-0001 dataset, MRI sequences were matched to the following standardized categories: T1-weighted (T1), T2-weighted (T2), T2 fluid-attenuated inversion recovery imaging (FLAIR), diffusion-weighted imaging (DWI), functional MRI (fMRI), susceptibility-weighted imaging (SWI), perfusion-weighted imaging (PWI), magnetic resonance angiography (MRA), localizer, other uncommon sequences, and unknown (ie, unable to classify). Parameter thresholds used for sequence identification are provided in the Supplement (Table S1). To improve sequence identification accuracy, an additional keyword-based matching step was applied using the scanner-generated sequence description. Though applied to all data, only sequences retaining informative descriptions after de-identification could be matched with keywords. This step captures terms indicative of less common sequences, including variants of SWI, PWI, and MRA, as well as b-value-specific descriptors in DWI. If there was a discrepancy, the keyword-based result was used to override the previous classification. This additional step was applied only to less common sequences, which often use acquisition parameters similar to common structural sequences, while relying on distinguishing features (e.g., bval for DWI). If these features are missing due to data incompleteness or de-identification, the sequences can be misclassified based on the broader parameter ranges of structural sequences. Keyword-based identification can therefore improve classification accuracy.

To estimate the accuracy of our sequence identification process, trained staff manually annotated a “ground truth” cohort of 100 sessions for each dataset (I-0001 and I-0004), which captured a broad range of Study Description (0008, 1030) tags across both sites. For each dataset, the ground truth cohort consisted of 2randomly sampled sessions from each of the top 20 most frequent Study Description tags and 60 additional sessions randomly sampled from the remaining tags. Overall sequence classification accuracy was reported for each dataset, along with the sensitivity and precision for T1, T2, and FLAIR classifications, for all original scans (non-derived according to Image Type) that were not manually labeled as “unknown”.

The custom code for sequence identification is available on Github (https://github.com/bdsp-core/BigBrainImagingDatabase).

#### 3.3.2 Clinical Metadata Extraction

Clinical metadata extraction from unstructured clinical imaging reports was performed using Bio-Medical Large Language Model (LLM) through Bio-Medical-Llama-3-8B,^19^ a model fine-tuned specifically for processing information from unstructured clinical text, which allowed us to efficiently process large volumes of clinical text data. We applied a stepwise approach to extract structured clinical information from unstructured clinical reports, using continuous and categorical prompts (see Figure 1). Whereas continuous prompts allowed an open-ended answer with few constraints, categorical prompts asked the model to choose from a predefined set of answers. This dual-approach strategy enabled us to capture a broad spectrum of heterogeneous findings while ensuring robust standardization.

**Figure 1:**
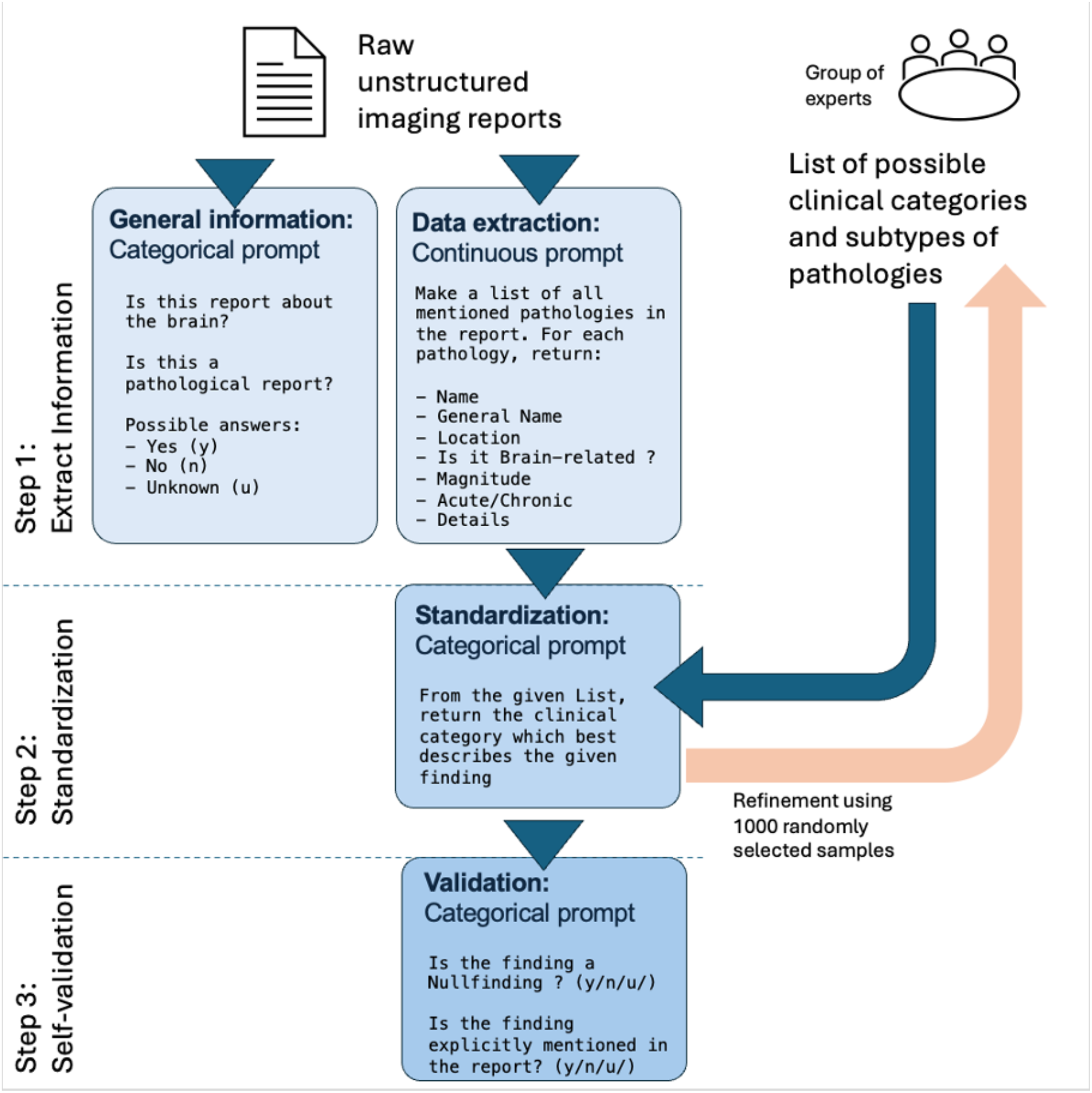
Workflow diagram of the LLM.

**Figure 2:**
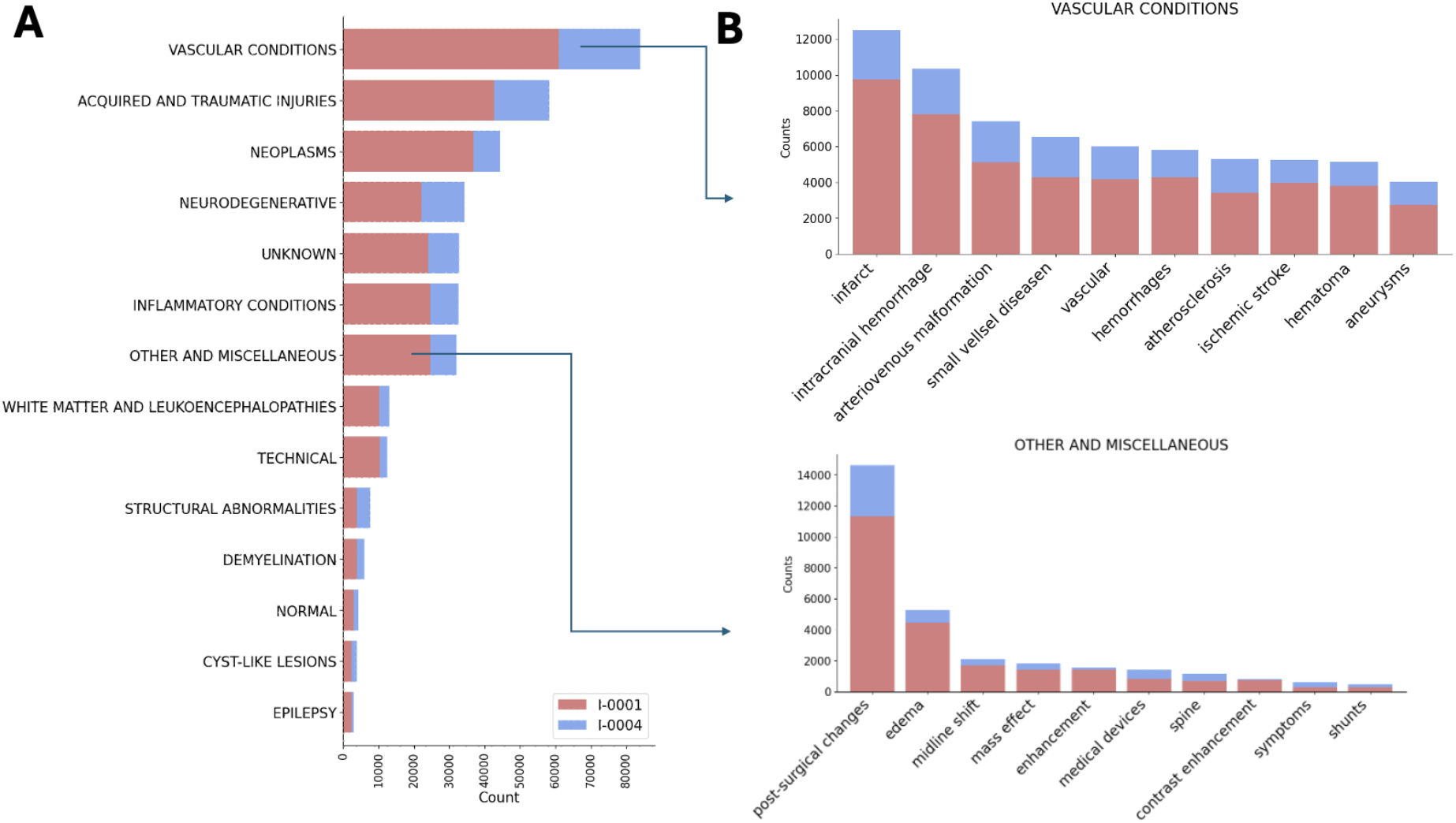
Brain-related findings.

In the first step (i.e., information extraction), the whole unstructured imaging report was given to the Bio-Medical LLM. The model was tasked with categorizing each report as either related to the brain or not, as well as identifying the presence or absence of any pathological condition in the report. The model responses were limited to ‘Yes’, ‘No’, or ‘Unknown’. In the second step, the model was prompted to extract and list all mentioned pathologies in the report, prioritizing clinically important ones while ensuring that only explicitly mentioned findings were included. The prompt involved generating a JSON-formatted output with detailed information for each finding, including 1) type of pathology, 2) general clinical term, 3) location, 4) whether the pathology is brain-related, 4) magnitude of the finding (for example size, severity, progression), and 5) whether it is acute, subacute or chronic.

In the third step (i.e., standardization), we aimed to assign standardized clinical categories to the lists of extracted information. Based on a list of over 300,000 findings, experienced neurologists (MBW, AL, SZ, PH, SM, JAK) iteratively reduced these to a list of 10 overarching clinical categories with 150 subcategories (see supplementary material, Table S2). Using each finding individually, LLAMA was prompted to select the clinical category that best represents the given finding. The prompts and list of pathologies were adjusted through a series of dialogue-driven refinements with neurologists, evaluating model performance using 1000 randomly selected samples and multiple clinicians (see Limitations).

In the last step (i.e., Self-validation), the model was asked whether each of the pathologies it had extracted was explicitly mentioned in the corresponding imaging report. This step aimed to identify over-interpretations of the findings or pathologies that were hallucinated by the model.

The overall performance of the approach was evaluated by a board-certified neurologist (PH) using 1000 randomly selected reports. Due to the nature of LLMs and the infeasibility of manually verifying results in almost 2 million brain scans, however, we cannot guarantee the accuracy or completeness of the extracted information. The standardized metadata should not be treated as the clinical ground truth but instead should serve as an entry point for the user, to facilitate access and navigation of the dataset and identification of potential clinical subgroups of interest. De-identified clinical reports are available in the database alongside the extracted metadata and should be reviewed by the user for confirmation. Clinical metadata should always be interpreted in accordance with the respective full-text clinical report.

## 4 Data Record

### 4.1 Dataset Storage and Organization

The BIND dataset is published as a part of the Brain Data Science Platform (https://bdsp.io/), a large collection of open-source clinical datasets for brain research. A data catalogue is provided in the supplemental material. BDSP repositories include the Human Sleep Project (HSP); the I-CARE (International Cardiac Arrest REsearch consortium), and others. On November 21, 2024, the NIH recognized BDSP as an approved data-sharing repository, aligning with the NIH’s DMS policy and underscoring its commitment to open science and collaborative neuroscience research. Data storage within BDSP is sponsored by the Amazon Web Services (AWS) Open Data Sponsorship Program, enabling secure, scalable, and free dataset access and sharing. For more information, visit bdsp.io (https://bdsp.io/) or explore BDSP resources (https://bdsp.io/content/).

The BIND dataset is organized in a standardized hierarchical folder structure, using Brain Imaging Data Structure (BIDS) format.^20^ Each site is represented by a top-level directory containing subdirectories for individual patients, followed by sessions and types of neuroimaigng data (anat/dwi/func/swi/etc.). Demographic information and clinical notes are provided in the non-BIDS counterpart for each patient and each session. A comprehensive table of all clinical metadata, data availability, and demographics is provided at the level of each site. Additional directories are provided with the original de-identified unstructured imaging reports. Of the patients included in the BIND dataset, 74.9% are also part of the Harvard Electroencephalography Database.^16^

Imaging files are provided in NIFTI format. Accompanying metadata are provided as .json or .csv files. Imaging reports are available as .txt files.

### 4.2 Demographics

#### 4.2.1 MGB Dataset (Subset I-0001)

Table 1 summarizes the demographic information for the BIND dataset. The subset from Site I-0001 (MGB) contains 81,297 clinical encounters from 28,438 individuals (13,866 male, 14,327 female, 245 unknown). At the earliest encounter within the dataset, the patients’ average age was 55.6 ± 21.8 years, ranging from 1 year to 106 years (Table 1). Individuals in the dataset self-identified with over 40 racial and ethnic attributions. A majority of subjects (72.53%) identified as White, followed by 7.4% Black or African American and 3.06% Asian. Nine percent of individuals identified with other racial attributes, including Native American or Alaska Native, Native Hawaiian or Other Pacific Islander. For 8.01% of patients, the race category is unknown.

**Table 1:** Demographic information, Table only displays a selection of common categories; additional categories available in the dataset demographic table.

|  |  | I-0004 | I-0001 |
| --- | --- | --- | --- |
| **Counts** | Subjects [N] | 10.504 | 28.438 |
|  | Sessions [N] | 27.440 | 81.297 |
| **Sex** | Female | 5085 (48.41%) | 14311 (50.32 %) |
|  | Male | 5418 (51.58 %) | 13845 (48.68 %) |
|  | Unknown | 1 (0.01%) | 282 (0.99 %) |
| **Age** | 0-5 years [N(%)] | 814 (7.75)% | 638 (2.24)% |
|  | 5-10 years [N(%)] | 460 (4.38)% | 314 (1.1)% |
|  | 10-15 years [N(%)] | 497 (4.73)% | 328 (1.15)% |
|  | 15-20 years [N(%)] | 452 (4.3)% | 522 (1.84)% |
|  | 20-30 years [N(%)] | 664 (6.32)% | 2208 (7.76)% |
|  | 30-40 years [N(%)] | 872 (8.3)% | 2326 (8.18)% |
|  | 40-50 years [N(%)] | 1251 (11.91)% | 2500 (8.79)% |
|  | 50-60 years [N(%)] | 1580 (15.04)% | 3893 (13.69)% |
|  | 60-70 years [N(%)] | 1773 (16.88)% | 5172 (18.19)% |
|  | 70-80 years [N(%)] | 1488 (14.17)% | 4735 (16.65)% |
|  | 80-90 years [N(%)] | 581 (5.53)% | 2366 (8.32)% |
|  | over 90 years [N(%)] | 70 (0.67)% | 463 (1.63)% |
| **Race** | White | 6006 (57.18 %) | 20606 (72.46 %) |
|  | Asian | 1553 (14.78 %) | 869 (3.06%) |
|  | Black | 444 (4.23 %) | 2103 (7.4 %) |
|  | Native American | 51 (0.49%) |  |
|  | Pacific Islander | 131 (1.25 %) |  |
|  | Unknown | 349 (3.32 %) | 2277 (8.01 %) |
|  | Other | 1970 (18.75 %) | 2556 (9.00%) |

Metadata tables with demographic information are provided in the dataset. Alongside the information described above, the demographic metadata contain the primary cause, date, and age at death (if recorded), marital status, occupation, language spoken, religion, veteran status and education level.

#### 4.2.2 Stanford Dataset (Subset I-0004)

A summary of all demographic information can be found in Table 1. The subset I-0004 (Stanford) contains 27,440 clinical encounters from 10,504 patients (5085 female, 5418 male, 1 unknown). At the earliest encounter within the dataset, the patients’ average age was 46.82 ± 25.16 years, ranging from newborn to 101 years (Table 1). Patients in this dataset identified as White (57.18%), Asian (14.78%), Black (4.23%), Pacific Islander (1.25%), Native American (0.49%) and Other (18.75%). For 3.32% of subjects, the racial identity is unknown.

Metadata tables with all demographic information are provided in the dataset. Alongside the information described above, the demographic metadata also contain the date and age at death (if recorded), marital status, occupation, language spoken, height, weight, and body mass index. In addition, metadata contains information on smoking history and alcohol use. Whereas Table 1 only displays general information on sex and race, more detailed information on race, ethnicity, and ethnic background can be found in the metadata.

### 4.3 Imaging Modalities and Sequences

Available scan modalities are summarized in Table 2. In summary, the BIND dataset comprises 1,791,885 clinical images, including 1,723,699 (96.19%) MRI; 54,137 (3.02%) CT; 5,093 (0.28%) PET scans; and 526 (0.03%) SPECT scans. Scanning sequences were chosen in a clinical context and are thus not standardized. Computationally-identified sequences are summarized in Table 3 (see Section 3.3 Computational processing, 3.3.1 Sequence names).

**Table 2:**
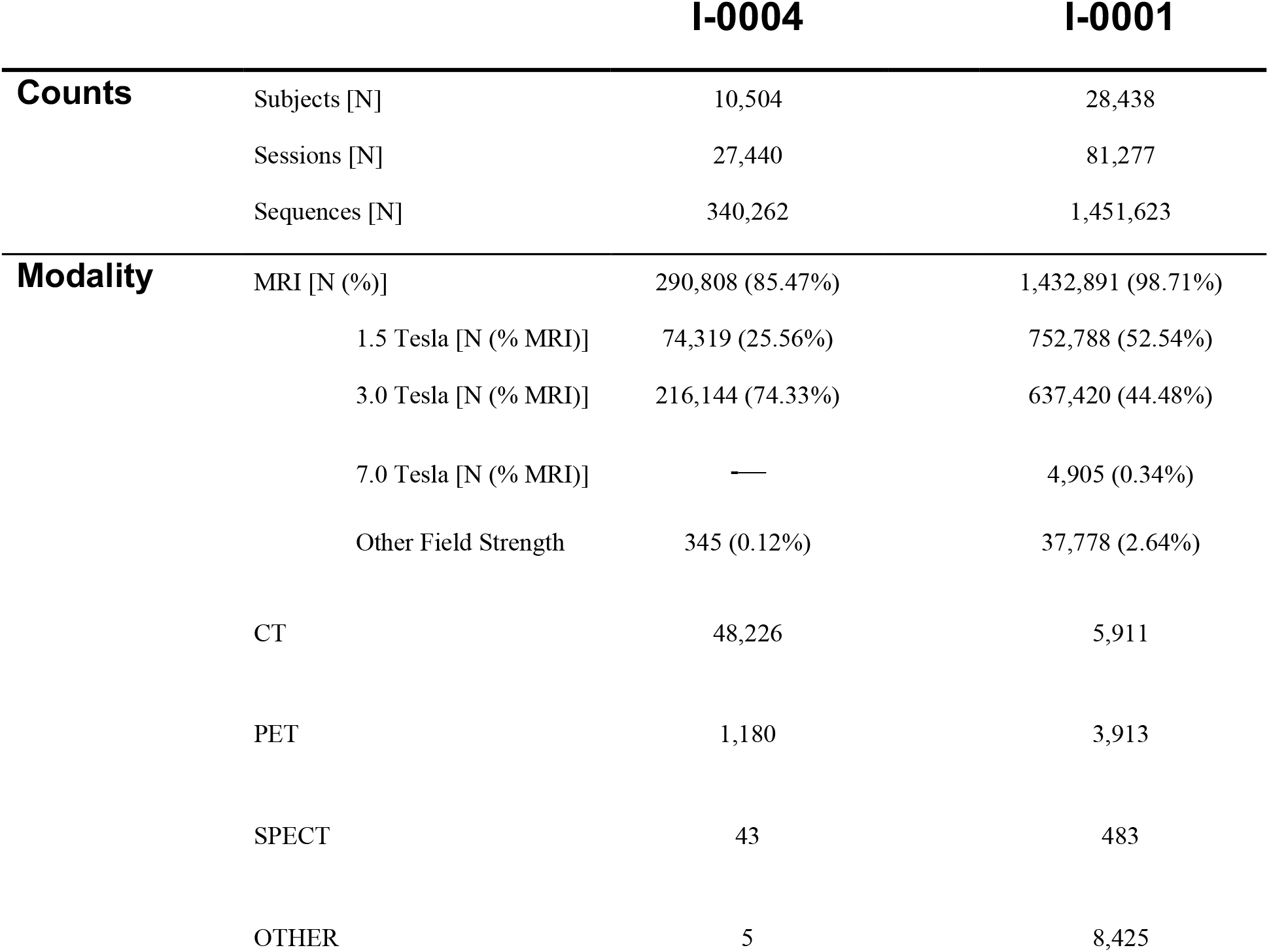
Scan modalities.

|  |  | I-0004 | I-0001 |
| --- | --- | --- | --- |
| Counts | Subjects [N] | 10,504 | 28,438 |
|  | Sessions [N] | 27,440 | 81,277 |
|  | Sequences [N] | 340,262 | 1,451,623 |
| Modality | MRI [N (%)] | 290,808 (85.47%) | 1,432,891 (98.71%) |
|  | 1.5 Tesla [N (% MRI)] | 74,319 (25.56%) | 752,788 (52.54%) |
|  | 3.0 Tesla [N (% MRI)] | 216,144 (74.33%) | 637,420 (44.48%) |
|  | 7.0 Tesla [N (% MRI)] | — | 4,905 (0.34%) |
|  | Other Field Strength | 345 (0.12%) | 37,778 (2.64%) |
|  | CT | 48,226 | 5,911 |
|  | PET | 1,180 | 3,913 |
|  | SPECT | 43 | 483 |
|  | OTHER | 5 | 8,425 |

**Table 3:** Computationally-identified MRI sequence names.

|  |  | I-0004 | I-0001 |
| --- | --- | --- | --- |
| Sequence | T1 | 71,931 | 366,228 |
|  | T2 | 56,307 | 110,083 |
|  | (T2) FLAIR | 18,728 | 101,857 |
|  | DWI (bval>0) | 68,730 (32,801) | 297,927 (13,277) |
|  | fMRI | 4 | 6,051 |
|  | SWI | 4,614 | 161,137 |
|  | PWI | 15,313 | 31,752 |
|  | MRA | 22,445 | 174,357 |
|  | Other (including Localizer) | 12,993 | 161,473 |
|  | Unable to identify [N (%)] | 19,743 (6.79%) | 22,026 (1.54%) |

### 4.4 Clinical Metadata

For 96.54% (I-0001: 100%, I-0004: 95.37%) of sessions, the session-specific clinical report is available and provided in the database. Using the LLM, 84,960 reports (87.31%) were determined to involve brain-related findings (see data usage note). Of available reports, 17,209 reports (17.69%) were identified as describing no pathology, 76,274 reports (78.39%) describe pathological findings, and 3,822 reports (3.93%) were categorized as unknown in terms of pathology (ie, based on those reports, the language model could not decide whether a pathology is present).

A standardized clinical metadata table is accessible to facilitate clinical data selection and interpretation (see Sections 3 Methods, 3.3.2 Clinical metadata extraction, and 6 Usage Notes/Data Availability). The clinical LLM extracted a total of 394,122 findings from all imaging reports (mean = 3.05 findings per report). For 318,726 (80.87%) of those findings, the model indicated a relation to the brain. Within those findings, 22.88% were categorized as a vascular condition, followed by 15.97% acquired and traumatic injuries, 12.17% neoplasms, 9.41% neurodegenerative, 8.95% inflammatory, 3.57% white matter conditions, 2.1% structural abnormalities, and 1.04% cyst-like lesions. Another 8.78% of data were labeled as “other” and “miscellaneous”, while 8.98% of findings could not be assigned a category. Of the total findings, 3.4% were related to technical specificities, artifacts, or signal alterations (Supplementary Table 2).

### 4.5 Limitations & Expected Future Updates

During the institutional de-identification process, some of the header information for the diffusion scans, namely the B values and B vectors, were overwritten. For the current dataset, approximately 12.57% of the diffusion scans have missing (overwritten) B values and B vectors. Future updates to the dataset will incorporate the B values and B vectors for all as well as expand the participating institutions for both the neurophysiology and neuroimaging datasets.

## 5 Technical Validation

### 5.1 MRI Sequence Name Identification Accuracy Estimation

MGB (I-0001) dataset: From a random sample of 100 imaging sessions, 1,039 sequences were included from the ground truth cohort, as they were not derived images and were not manually labeled as “unknown” sequences. The overall classification accuracy was 0.9471. Sensitivity and precision were: T1 (sensitivity: 0.8869, precision: 0.9960), T2 (sensitivity: 0.9793, precision: 0.9530), and FLAIR (sensitivity: 1.0, precision: 1.0).

Stanford (I-0004 dataset): From a random sample of 100 imaging sessions, 1,191 sequences met the inclusion criteria as above. The overall classification accuracy was 0.8455. Among the most clinically relevant categories, sensitivity and precision were: T1 (sensitivity: 0.9099, precision: 0.8189), T2 (sensitivity: 0.9711, precision: 0.8131), and FLAIR (sensitivity: 1.0, precision: 1.0). Additional confusion matrixes are included in the supplement (Figure S1). These results support the reliability of our tool in accurately identifying MRI sequence types across multi-site, heterogeneous datasets.

## 6 Usage Notes

### 6.1 How to Access the Data

Data access is provided via the Brain Data Science Platform. Data access instructions and security protocols are provided on **bdsp.io** (https://bdsp.io//about/howto_accessdata/). To set up data access, an active AWS account with AWS account ID is required and must be provided in the BDSP profile settings. To ensure alignment with strict data security protocols, researchers are required to sign a Data Use Agreement and provide proof of passing CITI Training certification tests. After approval of the application, data can be accessed through the AWS Command Line Interface, using AWS Access Keys. BDSP allows users to list directories, download files, and copy entire folders to their local systems, or to work with the data in the cloud.

### 6.2 Use of Clinical Metadata

Due to inherent limitations of large language models and the absence of human-curated metadata, we cannot guarantee the accuracy or completeness of the information extracted from the dataset. Instead, the standardized metadata should be viewed as a preliminary contextual framework, enabling users to navigate and search the dataset efficiently while identifying clinically-relevant subgroups. Full-text de-identified clinical reports are available within the database alongside the extracted metadata, serving as the authoritative source for interpretation and validation of clinical findings by the users. Clinical metadata should thus always be interpreted by the user in accordance with the respective full-text clinical report.

### 6.3 Use of Diffusion Images

Approximately 13% of the diffusion images are missing the corresponding B values and B vectors. As described above, this occurred during the robust institutional de-identification step, which overwrote these portions of the DICOM headers (Table 3). A table indicating the cases with this missing information is provided in the database. A future update to the database is expected to add the remaining B values and B vectors files to assist with diffusion processing.

### 6.4 Imaging Quality

The BIND dataset consists of unprocessed clinical imaging studies in NIfTI format, which were acquired without prior signal quality assessment or preprocessing. As a result, scanning quality, resolution, and levels of artifact variability can significantly impact image interpretation. Visual assessment of the images for identified subgroups of interest is recommended prior to analysis.

### 6.5 Sequence Identification

Despite the overall high accuracy of our sequence identification, several challenges and limitations remain. The heterogeneity of clinical imaging data collected across sites, including scans acquired in external institutions or using non-standardized protocols, introduces variability that may lead to misclassification. Differences in scanner vendors and field strengths; missing sequence descriptions due to the de-identification process; and the inclusion of research or experimental sequences add further complexity to labeling the sequences. For instance, localizers were occasionally identified as T1- or T2-weighted images due to site-specific variations and overlapping parameter values. Moreover, the I-0004 dataset included a substantial number of neonatal scans, which may have contributed to errors in sequence classification. Finally, the inclusion of non-brain images and uncommon sequence types in the dataset also likely contributed to classification error. These limitations highlight the importance of continued refinement of the sequence identification tool for future applications.

## Data Availability

The manuscript describes the The Brain Imaging and Neurophysiology Database (BIND) published at https://bdsp.io/content/n1vba1x5qt62frfjem65/1.0/

https://bdsp.io/

## 7 Data Availability

The BIND dataset is published as a part of the Brain Data Science Platform (https://bdsp.io/) and is available at (https://bdsp.io/content/n1vba1x5qt62frfjem65/1.0/).^21^ The corresponding EEG and PSG data were previously published by our group and can be found in the ‘Harvard Electroencephalography Database’ (https://bdsp.io/content/harvard-eeg-db/4.1/)^16,22^ and ‘The Human Sleep Project’ (https://bdsp.io/content/hsp/2.0/) ^23^, respectively. For more information on data access explore BDSP resources (https://bdsp.io/content/). Individuals seeking access to the data are required to sign a mandatory Data Use Agreement, which includes strict terms and conditions, and to provide proof of CITI training (https://about.citiprogram.org/) which can be freely accessed through the BDSP website. These agreements prohibit attempting to reidentify individual records or further sharing of the data. For more information, visit bdsp.io (https://bdsp.io/).

## 8 Code Availability

Custom code used for the LLM metadata extraction and sequence identification is available on Github (https://github.com/bdsp-core/BigBrainImagingDatabase). The directory also provides code to systematically search the database for a specific modality or finding.

## 9 Acknowledgements, Author Contributions & Competing Interests

This work was supported by grants from the NIH (RF1AG064312, RF1NS120947, R01AG073410, R01HL161253, R01NS126282, R01AG073598, R01NS131347, R01NS130119, R01NS131347). Dr. Westover is a co-founder, scientific advisor, consultant to, and has personal equity interest in Beacon Biosignals. SSM is supported by the National Institute of Mental Health [R01MH131194, R01MH134823], the Claflin Distinguished Scholar award [Massachusetts General Hospital]. ADL has served as a consultant for Neurona Therapeutics, and the institution of ADL has received research funding from Neurona Therapeutics and Sage Therapeutics. Dr. Silvers is employed by and has personal equity interest in AWS.

**Writing - Original Draft:** C. Maschke, P. Hadar, Y. Zhang; **Writing - Review & Editing:** all authors; **Software – information from clinical reports:** C. Maschke, P. Hadar; **Software – sequence identification:** Y. Zhang; **Software – image processing:** Y. Zhang, J. Li; **Data Curation:** G. Ganjoo, J. Li, A. Gupta, M. Ghanta, B. Nearing; **Funding acquisition:** A. Hoopes, R. Thomas, J. A. Kim, S. S. Mukerji, A. Dalca, S. Zafar, A. D. Lam, E. Mignot, M Brandon Westover, C. Tsien Silvers, B. Gunapati; **Conceptualization and Supervision:** C. Tsien Silvers, B. Gunapati, C. Maschke, P. Hadar, Y. Zhang, A. Hoopes, A. Guazzo, R. Thomas, J. A. Kim, S. S. Mukerji, A. Dalca, S. Zafar, A. D. Lam, E. Mignot, M B. Westover

