## supplementary material for "The Brain Imaging and Neurophysiology Database: BINDing multimodal neural data into a large-scale repository"

[7] Amazon Web Services

[8] Yale University, New Haven, CT, USA

This document contains:

- Supplemental Table 1-2
- Supplemental Figure 1
- Data Catalogue

**Supplemental Table 1:** MRI sequence identification parameters (TE, TR, and TI in ms)

| | Scanning Sequence | TE | TR | TI | Other Parameters |
|---|---|---|---|---|---|
| **T1** | Spin Echo | <= 25 | <= 1,500 | 0 or NaN | |
| | | | >= 1,000 | 400 - 1,800 | |
| | Gradient Echo | | 1,000 - 5,000 | 300 - 3,000 | MR Acquisition Type: 3D; Sequence Variant includes "MP" |
| | | < 10 | <= 500 | > 0 | Sequence Variant includes "SP" |
| **T2** | Spin Echo | >= 30 | >= 800 | 0 or NaN | |
| | | | >= 1,000 | <= 400 | |
| **FLAIR** | Gradient Echo | >= 10 | 120 - 1,600 | | |
| | Spin Echo | >= 60 | >= 2000 | >= 1,200 | |
| **DWI** | Echo Planar | >= 40 | >= 1,800 | | Diffusion B Value > 0 * |
| **fMRI** | Echo Planar | 25 - 50 | 1,000 - 3,000 | | Echo Train Length >= 30 |
| **SWI** | Echo Planar | 20 - 40 | <= 100 | | |
| | Gradient Echo | < 55 | < 120 | | MR Acquisition Type: 3D; Flip Angle <= 40° |
| **PWI** | Echo Planar | < 60 | > 1,000 | | Echo Train Length >= 1 |
| **MRA** | Gradient Echo | <= 9 | <= 40 | | |

**Supplemental Table 2:** Clinical categories

| General Clinical Category | Subcategories |
|---|---|
| **VASCULAR CONDITIONS** | |
| | Amyloid angiopathy |
| | Aneurysms |
| | Arteriovenous malformation |
| | Atherosclerosis |
| | Capillary telangiectasia |
| | Carotid-cavernous fistula |
| | Cavernous malformation |
| | Developmental venous anomaly |
| | Dural arteriovenous fistula |
| | Hemorrhages |
| | hypoxic ischemic injury |
| | Intracranial hemorrhage |
| | ischemic stroke |
| | Moyamoya disease |
| | Pseudoaneurysm |
| | Reversible Cerebral Vasoconstriction Syndrome |
| | Small vessel ischemic disease |
| | Thrombosis |
| | Vasculitis |
| | Vaso-occlusive disease |
| | Veno-occlusive disease |
| | Venous angioma |
| | Venous sinus thrombosis |
| | Venous stasis |
| **NEOPLASMS** | |
| | Embryonal Tumors |
| | Glial Tumors |
| | Hematopoietic and Lymphoid Neoplasms |
| | Meningeal Tumors |
| | Metastases |
| | Other or Miscellaneous Tumors |
| | Schwannomas |
| | Other Nerve Sheath Tumors |
| | Sellar, Parasellar, and Suprasellar Tumors |

| | |
|---|---|
| **NEURODEGENERATIVE** | Alzheimer's disease<br>Amyotrophic lateral sclerosis (ALS)<br>Atrophy and Sulci Enlargement<br>Corticobasal Degeneration<br>Creutzfeldt Jakob disease (CJD)<br>Frontotemporal dementia (Pick's disease)<br>Huntington's disease<br>Multiple System Atrophy (MSA)<br>Normal Pressure Hydrocephalus (NPH)<br>Parkinson's disease<br>Progressive Supranuclear Palsy (PSP)<br>Vascular dementia |
| **DEMYELINATION** | Multiple Sclerosis (MS)<br>Neuromyelitis Optica (NMO, Devic's)<br>MOG antibody disease<br>Acute Disseminated Encephalomyelitis (ADEM) |
| **STRUCTURAL ABNORMALITIES** | Agenesis or hypoplasia (of the corpus callosum)<br>Basilar invagination, Platybasia<br>Chiari malformations<br>Craniocervical junction anomalies<br>Cortical dysplasia<br>Dandy-Walker complex<br>Empty sella turcica<br>Encephalocele<br>Heterotopia<br>Holoprosencephaly<br>Lissencephaly<br>Mega cisterna magna<br>Neurofibromatosis (NF)<br>Polymicrogyria<br>Schizencephaly<br>Sturge-Weber syndrome<br>Tethered cord<br>Tuberous sclerosis<br>Von Hippel–Lindau (VHL) |
| **CYST-LIKE LESIONS** | Arachnoid cyst<br>Colloid cyst<br>Dermoid cyst<br>Epidermoid cyst<br>Mucous retention cyst<br>Pineal cyst<br>Porencephalic cyst<br>Pseudomeningocele<br>Rathke's cleft cyst<br>Subependymal cyst |
| **ACQUIRED and TRAUMATIC INJURIES** | Contusion (coup-contrecoup injuries)<br>Hematoma<br>Herniation |

| | |
|---|---|
| | Hydrocephalus<br>Idiopathic Intracranial Hypertension (IIH)<br>Intracranial hemorrhages<br>Leptomeningeal enhancement<br>Mesial temporal sclerosis (often in epilepsy)<br>Normal Pressure Hydrocephalus (NPH)<br>Papilledema<br>Pituitary apoplexy<br>Pneumocephalus<br>Posterior Reversible Encephalopathy Syndrome (PRES)<br>Seizure-related changes<br>Shear injury / Diffuse axonal injury (DAI)<br>Skull fracture<br>Toxic-metabolic injury<br>Traumatic brain injury (TBI) |
| **INFLAMMATORY CONDITIONS** | Abscess<br>Autoimmune conditions<br>Behçet's disease (Neuro-Behçet's)<br>Cerebritis<br>Creutzfeldt Jakob disease (infectious prion)<br>Empyema<br>Encephalitis<br>Granulomatous disease<br>HIV encephalopathy<br>Labyrinthitis<br>Mastoiditis<br>Meningitis<br>Neurocysticercosis<br>Prion diseases (rare variants)<br>Progressive Multifocal Leukoencephalopathy<br>Sarcoidosis<br>Sinus disease<br>Susac's syndrome<br>Toxoplasmosis<br>Ventriculitis |
| **WHITE MATTER AND LEUKOENCEPHALOPATHIES** | Binswanger's disease (subcortical leukoencephalopathy)<br>Cerebral Autosomal Dominant Arteriopathy with Subcortical Infarcts and Leukoencephalopathy (CADASIL)<br>Central pontine myelinolysis (osmotic demyelination)<br>Leukoencephalopathy or white matter disease<br>Leukomalacia<br>Metabolic leukodystrophies<br>Radiation-induced leukoencephalopathy<br>Toxic leukoencephalopathy |
| **OTHER AND MISCELLANEOUS** | Artifact<br>Cardiac<br>Contrast enhancement<br>Edema<br>Hepatic<br>Lab measurement<br>Mass effect<br>Medical Devices<br>Midline shift<br>non-diagnostic finding<br>Post-Surgical Changes |

| | |
|---|---|
| | Proptosis<br>Pulmonary<br>Scan Type<br>Shunts<br>Signal alterations<br>Spine<br>Spleen<br>symptoms<br>thyroid |

**Supplemental Figure 1:** Confusion matrixes for MRI sequence identification. (A) I-0001; (B) I-0004.

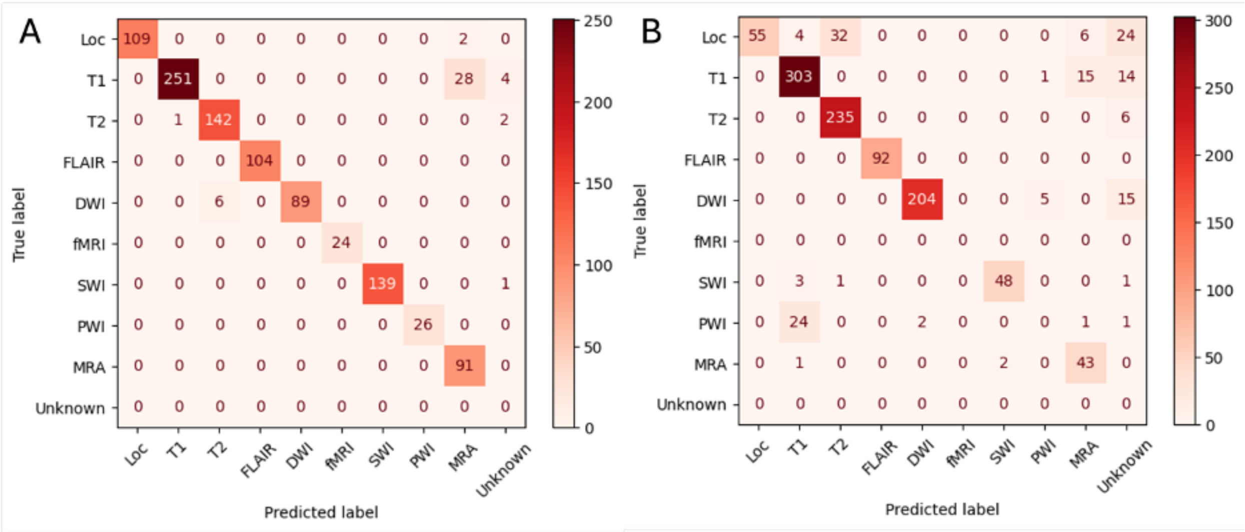

### Data Catalogue

#### MGB – I0001 Metadata

##### Total subject count:

- 28 257 subjects

BIDS data directory:

- 27 711 subject folders

  (from which 27 684 also in non-BIDS)

Non-BIDS data directory:

- 28 230 subject folders

  (from which 27 684 also in BIDS)

  +181

##### Demographics.csv

**Rows:** 81297 [Sessions from 28438 Subjects]

**Columns:** 14

1. 'Bdsp_patient_id'
2. 'Session_id'
3. 'ShiftedDateOfBirth'
4. 'ShiftedStudyDate'

5. 'PatientRace'
6. 'EthnicGroup'
7. 'MaritalStatus'
8. 'Religion'
9. 'Language'
10. 'VeteranStatus'
11. 'Sex'
12. 'EducationLevel'
13. 'GenderIdentity',
14. 'SexAssignedAtBirth'

**Reports.csv**

**Rows:** 81297 [Sessions from 28438 Subjects]

**Columns:** 4

1. 'Bdsp_patient_id'
2. 'Session_id'
3. 'Type': Type of session
4. 'Report_txt': Anonymized open-text reports, not standardized

### Stanford – I0004 Metadata

##### Total subject count:

- 10 504 subjects

BIDS data directory:

- 8 593 subject folders

  (from which 8 158 also in non-BIDS)

Non-BIDS data directory:

- 10 069 subject folders

(from which 8 158 also in BIDS)

#### Demographics.csv

**Rows:** 27440 [Sessions from 10.504 subjects]

**Columns:** 24

1. 'Bdsp_patient_id'
2. 'Session_number'
3. 'sex'
4. 'Shifted_date_of_birth'
5. 'Shifted_encounterdate'
6. 'age'
7. 'shifted_date_of_death'
8. 'age_at_death'
9. 'race'
10. 'ethnicity'
11. 'marital_status'
12. 'Occupation'
13. 'language'
14. 'shifted_encounter_date'
15. 'recent_height_cm'
16. 'recent_weight_kg'
17. 'recent_bmi'
18. 'smoking_hx'
19. 'shifted_smoking_started'
20. 'shifted_smoking_quit'
21. 'shifted_smoking_history_taken'
22. 'alcohol_use'
23. 'deceased'
24. 'Ethnic_background'

#### Reports.csv

**Rows:** 27440 [Sessions from 10.504 Subjects]

**Columns:** 4

1. 'Bdsp_patient_id'
2. 'Session_id'
3. 'Report_txt': Anonymized open-text reports, not standardized
4. 'Type': Type of session

#### **Findings.csv** (available for both sites)

**Rows:** N [N Findings from all Subjects]

MGB: 279911 findings from 27076 subjects

STANFORD: 114211 findings from 10488 Subjects

**Columns:** 17 [Features, see below]

**Features:**

**General Features (column 1-5)**

1. **Site:** MGB or STANFORD
2. **Bdsp_patient_id**
3. **Session_id**
4. **Counts:** Number of finding counted per scan
5. **Abnormality:** Is this a pathological report? Answer 'Yes', 'No' or 'Unknown'.
6. **Brain:** Is this report about the brain? Answer 'Yes', 'No' or 'Unknown'

**Answers from Step 1 model (column 5-12)**

Open question: Make a list of all mentioned pathologies in the report, start with the most clinically important one. Do not repeat pathologies. Only include pathologies which are explicitly mentioned in the report. Return a list of pathologies in JSON format. Each finding should have a 'Name', 'General Name', 'Location', 'Brain-related', 'Magnitude','Acute/Chronic'

7. **Name:** Name of the pathology
8. **General Name:** General name of the pathology
9. **Location:** Location of the pathology
10. **Brain-related:** Is the pathology Brain related
11. **Magnitude:** of the pathology
12. **Acute/Chronic:** can be subacute as well

**Answers from Step 2 model (column 13-17)**

13. **Test:** Sanity check for previous model to detect hallucinations. Was the finding explicitly mentioned in the report? Please be cautious interpreting the finding if the answer is no. It can be a hallucination, however it can also be a clinical implication which was not explicitly mentioned in the report.
14. **Label_manual:** Using the general Name, we performed a manual labeling or cleaning of labels using simple rules, to manually standardize the names (ie. for example removing plural forms and synonyms).
15. **Nullfinding:** The model had issues with negations, so we asked an additional question, whether the given finding is a null finding or negative finding. Please be careful interpreting the finding if the answer is yes. This question aimed to identify findings with terms such as 'no evidence of…' or 'normal'.
16. **Label_LLAMA_Level2:** low-level clinical Category chosen by LLAMA (see supplementary Table 2). Categories are not exclusive. For some findings, more than one category applies.
17. **Label_LLAMA_Level1:** higher-level clinical Category chosen by LLAMA (see supplementary Table 2). Categories are not exclusive. For some findings, more than one category applies.
